# Identification of thresholds on population density for understanding transmission of COVID-19

**DOI:** 10.1101/2022.01.27.22269840

**Authors:** Yusuf Jamal, Mayank Gangwar, Moiz Usmani, Alison Adams, Chang-Yu Wu, Thanh Huong Nguyen, Rita Colwell, Antarpreet Jutla

**Affiliations:** GeoHLab, Department of Environmental Engineering Sciences, University of Florida, Gainesville, FL; Community and Environmental Sociology, University of Florida, Gainesville, Florida, FL; Department of Environmental Engineering Sciences, University of Florida, Gainesville, FL; Civil and Environmental Engineering, University of Illinois, Urbana, IL; University of Maryland Institute of Advanced Computer Studies, University of Maryland, College Park, MD

**Keywords:** threshold, population density, logistic regression, COVID-19

## Abstract

Pathways of transmission of coronavirus (COVID-19) disease in the human population are still emerging. However, empirical observations suggest that dense human settlements are the most adversely impacted, corroborating a broad consensus that human-to-human transmission is a key mechanism for the rapid spread of this disease. Here, using logistic regression techniques, estimates of threshold levels of population density were computed corresponding to the incidence in the human population. Regions with population densities greater than 3000 person per square mile in the United States have about 95% likelihood to get infected with COVID-19. Since case numbers of COVID-19 dynamically changed each day until November 30, 2020, ca. 4% of US counties were at 50% or higher risk of COVID-19 transmission. While threshold on population density is not the sole indicator for predictability of coronavirus in human population, yet it is one of the key variables on understanding and rethinking human settlement in urban landscapes.

**Plane language Summary:** Population density is certainly one of the key factors influencing the transmission of infectious diseases like COVID-19. It is approximated that in continental United States, population density of 1192 per square mile and higher presents 50% probability of getting infected with COVID-19.

**Key Points:**

- Based on data from the USA, the population density of 1192 persons per square mile represented a 50% or higher probability of risk of transmission of COVID-19.
- About 35 counties in the USA are at very high risk of transmission potential (95% or higher) for COVID-19.
- Analysis shows the vulnerability of urban towns to respiratory infectious disease

## 1. Introduction

Severe Acute Respiratory Syndrome caused by Coronavirus (SARS-CoV-2 thereafter) is a respiratory lung infection, and as of April 28, 2021, there have been more than 148 million (WHO COVID-19 Dashboard, https://covid19.who.int/) confirmed human cases in the world. The SARS-CoV-2 virus remains highly infectious and is circulating in the human population at an alarming rate with anticipated variants in near future. An emerging disciplinary consensus is that seasonal variation may lead to cyclical outbreaks in the human population(Carlson, Gomez, Bansal, & Ryan, 2020; Merow & Urban, 2020). As with all airborne respiratory infectious diseases, the transmission of SARS-CoV-2 is high in densely populated urban regions of the world (Cruickshank, 1939; Robinson, Stilianakis, & Drossinos, 2012). However, thresholds of population density relative to the outbreak of the disease in humans remains unknown. The relative importance of knowledge of threshold on population density with reference to infectious disease such as COVID-19 is important for the future of modern cities and urban landscapes in the USA, given about 71% of the population reside in urbanized areas with an average density of 2534 persons per square mile (https://www.census.gov/programs-surveys/geography/guidance/geo-areas/urban-rural/ua-facts.html).

Influenza transmission dynamics, which allow parallel comparison with COVID-19 transmission, depend on several socio-demographic factors (such as race, income level, education, and location), but population density remains a critical variable for controlling an outbreak of seasonal influenza (Atkinson & Wein, 2008; Merler & Ajelli, 2010). While the severity of airborne contagion cannot be attributed solely to population density(Li, Richmond, & Roehner, 2018), the knowledge of thresholds on population density can be helpful in understanding the spatial distribution with respect to the risk of disease(Chandra, Kassens-Noor, Kuljanin, & Vertalka, 2013; Grantz et al., 2016). Intuitively, high population density is concluded to favor contagion and vice-versa. However, non-uniform distribution of a population can yield inconclusive results (Li et al., 2018). While the significant association was reported between population density and transmissibility for the 1918 influenza pandemic in Chicago (Grantz et al., 2016), the average influenza attack rates decreased with increasing population density in Japan (Hoyle & Wickramasinghe, 1990). In the context of COVID-19, an analysis of Brazilian data suggests the general increase in COVID-19 cases was associated with highly populated regions(Pequeno et al., 2020). There is no study to date that provides an exploratory association of population density thresholds with COVID-19 cases in the continental USA. This study was undertaken to determine thresholds on population density that can be used to estimate the probable risk of infection from COVID-19. Estimation of a threshold population density would allow in the differentiation of low and high-risk regions and offer useful input for planning, designing, and targeting public health interventions. Also, identifying specific regions where greater surveillance is required to contain the disease would be enhanced and can be used to define the expansion of urbanized areas in the USA.

## 2. Methods

Daily incidence data for COVID-19 cases in the 3,107 mainland U.S. counties were obtained from the GitHub project (https://github.com/nytimes/covid-19-data) from March 15, 2020, to November 30, 2020. The time period was selected based on the non-availability of vaccines since vaccines will mask the limit on population densities. Data on the population densities of each U.S. county were obtained from the U.S. Census Bureau (2019) (https://www.census.gov/data/datasets/time-series/demo/popest/2010s-counties-total.html). Land area in square miles was obtained from the U.S. Census Bureau (https://www.census.gov/library/publications/2011/compendia/usa-counties-2011.html#LND). An ordinal logistic regression model was employed to study the dependency of COVID-19 cases (thereafter cases) on counties’ population density. It was assumed that the population densities remained constant during the study period, implying population mobility has minimal impact on population density. Further, the population was assumed to be uniformly distributed over the county. Since the number of cases differs widely over the county, intuitively, it is preferable to classify cases into a number of classes where each class explicitly implies a specific infection risk. To classify cases, we estimated the percentile of cumulative cases at an interval of 15 days beginning March 15, 2020, to November 30, 2020. Cumulative cases data were divided into three categories (or events): (a) low number of cases (up to 80th percentile); (b) medium number of cases (80th to 95th percentile); and (c) high number of cases (greater than 95th percentile). The 80^th^ percentile case on November 30, 2020 was 3817 (Table 1), which can still be considered relatively less compared to the number of cases in high-risk locations.

**Table 1:** Biweekly 80^th^ percentile cases of low cases

| Date | 80th percentile cases |
| --- | --- |
| March 15 2020 | 0 |
| March 31 2020 | 15 |
| April 15 2020 | 63 |
| April 30 2020 | 126 |
| May 15 2020 | 196 |
| May 31 2020 | 271 |
| June 15 2020 | 351 |
| June 30 2020 | 480 |
| July 15 2020 | 670 |
| July 31 2020 | 970 |
| August 15 2020 | 1175 |
| August 31 2020 | 1382 |
| September 14 2020 | 1564 |
| September 30 2020 | 1787 |
| October 15 2020 | 2069 |
| October 31 2020 | 2410 |
| November 15 2020 | 3036 |
| November 30 2020 | 3817 |

Moreover, a higher value for the log-likelihood (a statistical metric used for model selection) in Table 2, justifies the choice of the 80th percentile. In fact, the log-likelihood function was found to be better with increasing percentile, with the increase being less for the percentiles above the value of 80.

**Table 2:**
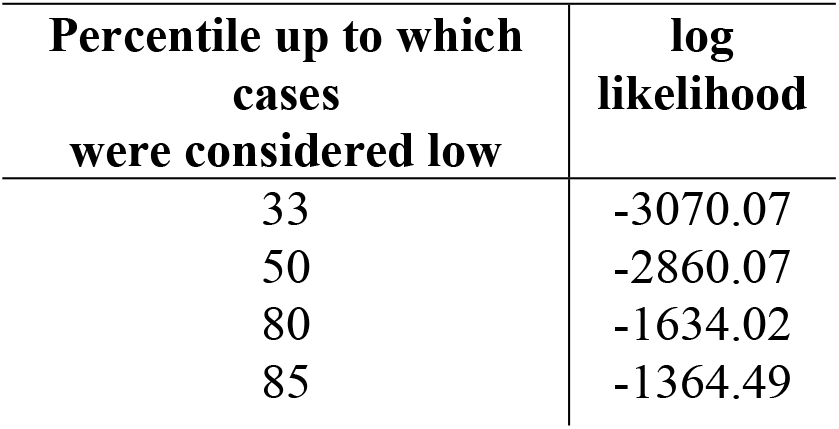
Variation in log-likelihood in COVID-19 cases

Thus, the ‘reasonable’ choice of percentiles for classification of cases will lead to overall similar model results without altering the final interpretation.. The following paragraph briefly explains the analysis approach. Relevant theory for the application of ordinal logistic regression is detailed in the supplementary section.

The predictor used is the county population density which is the county’s population per unit of the county’s land area. The response variable, which is ordinal in nature, is the cumulative case count classified into low, medium, and high (based on percentiles). Logit link function (details in supplementary section) was used to express the dependent variable as a linear function of the independent variables. Link function also relates the response (ordinal cases) to linear predictor (population density) and transforms the probabilities of ordinal response to the continuous scale [0,1]. The regression equations, thus, take the form as follows:

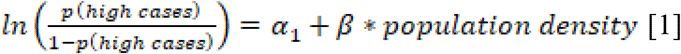

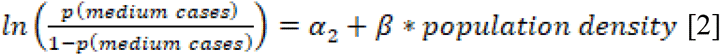

where *p(high cases)* and *p(medium cases)* are the probabilities of high and medium cases, respectively. Constants and coefficients in the equations (1, 2) were estimated using the maximum likelihood estimation methods. Since the total probabilities sum up to one, the probability of a low number of cases were estimated by subtracting the probability of high and medium cases.

## 3. Results and discussion

We start our analysis with results obtained from logistical regression models (Figure 1) showing three critical statistical metrics (Somers D, Goodman-Kruskal Gamma and Percent Concordant pairs).

**Figure 1:**
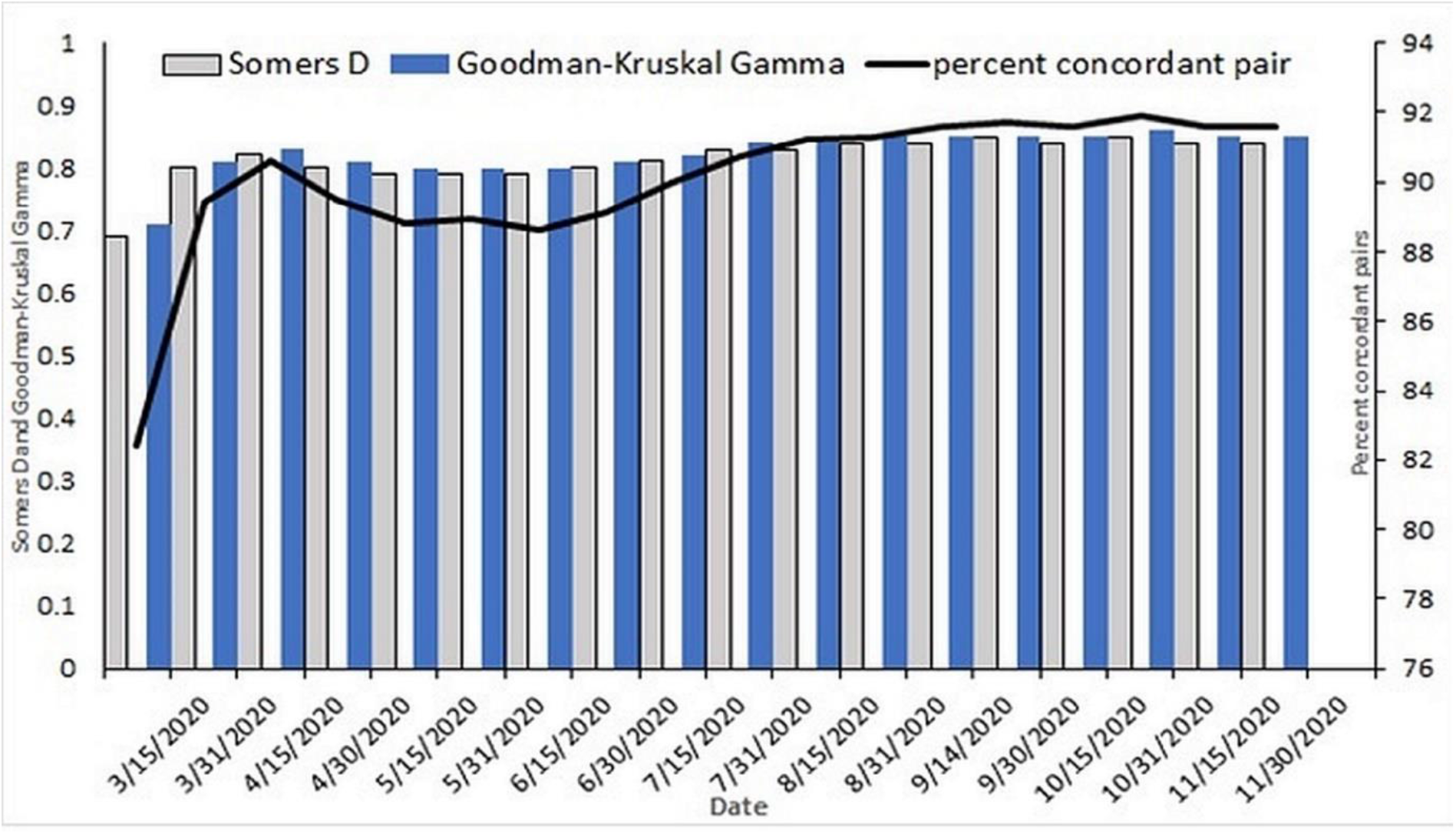
Performance of logistical regression model on a biweekly scale for the entire USA

High values of measures of association (> 82%), i.e., percentage of concordant pairs, Somers’s D and Goodman-Kruskal Gamma, signify that the performance of ordinal logistic model is satisfactorily. On average, model performance remained constant since start of collection of data on COVID-19 human cases. The p-values for each constant and predictor population density were less than 0.05 (not shown), thus establishing statistical significance. The p-value for the test of all slopes is zero (Table S1), which indicates the predictor population density has a statistically significant relationship with the response variable (COVID-19 cases). The deviance goodness of fit result (p>0.05), for all dates for which the model was run, showed adequate fit for the data, and the associated probabilities do not deviate significantly from observed values (detailed inference of model performance indicators for November 15, 2020 is presented in Table S1 in supplementary section). The model results for any one day should sufficiently explain the behavior of event probability (cases) with population density. The plots in Figure 2 show average probability of high, medium, and low number of cases.

**Figure 2:**
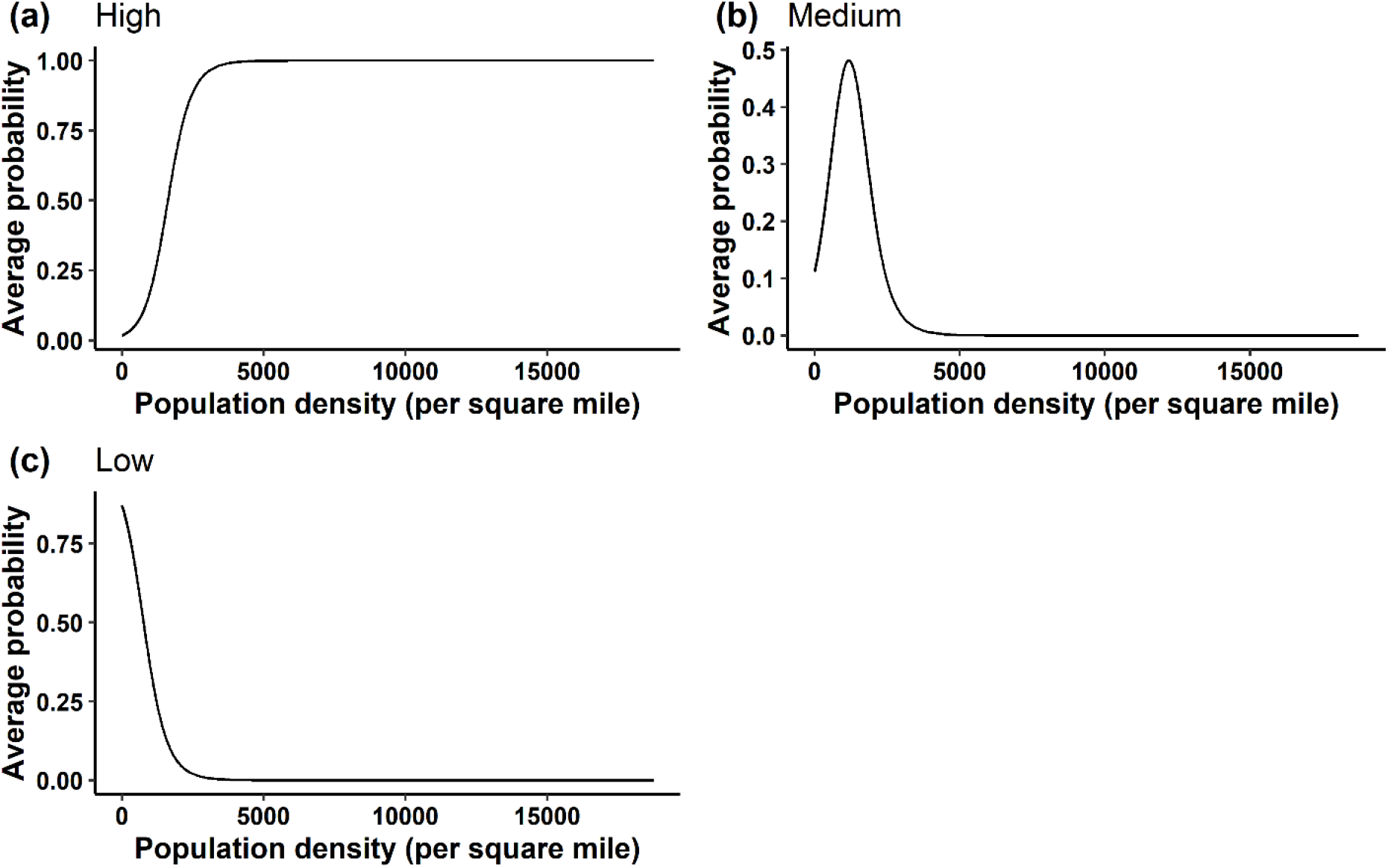
Average probability vs population density for (a) High (b) Medium (c) Low number of cases, from March 15, 2020, to November 30, 2020

Average probabilities were defined as the mean of the probabilities obtained from the ordinal logistic regression models for each of the eighteen bi-weekly cases (from March 15, 2020, to November 30, 2020). The monotonic nature of Figure 2(a) shows that with an increasing population density, the probability of a high number of cases increases, and vice versa. The probability rises steeply to a nearly constant value of 1 at a population density of ca. 5000 per square mile, suggesting larger population densities greater than this value were remarkably associated with the corresponding high number of COVID-19 cases. The implication is that the pronounced effect of high population density and a proportional number of cases was sufficient to establish population density as an important factor in transmission potential of this disease. The results suggest that in densely populated areas, it may be challenging to follow social distancing norms, thus an increased number of COVID-19 human cases were to be expected. Figure 2(c) shows the probability of a low number of cases, a trend opposite to that for high number of cases. The probability continuously decreases to a constant value close to zero, signifying that as population density increases, the chance of a low number of cases decreases. Figure 2(b) illustrates the medium number of cases with population density. The maximum probability of the medium number of cases is ca. 0.48, corresponding to the population density of ca. 1,190 per square mile. Thus, a decrease in population density from 1,190 per square mile decreases the probability of a medium number of cases as the probability of low number of cases increase. On the other hand, if population density beyond 1,190 per square mile, the probability for a medium number of cases decreases since probability of high number of cases increase thereafter. Population density of 1,190 people per square mile can be interpreted as a transition from low to high COVID-19 cases. Figure 3 illustrates changes in event probabilities over time and suggests that even low-density counties are likely to be more vulnerable as the probability of high number of COVID-19 cases for population density increases over time.

**Figure 3.**
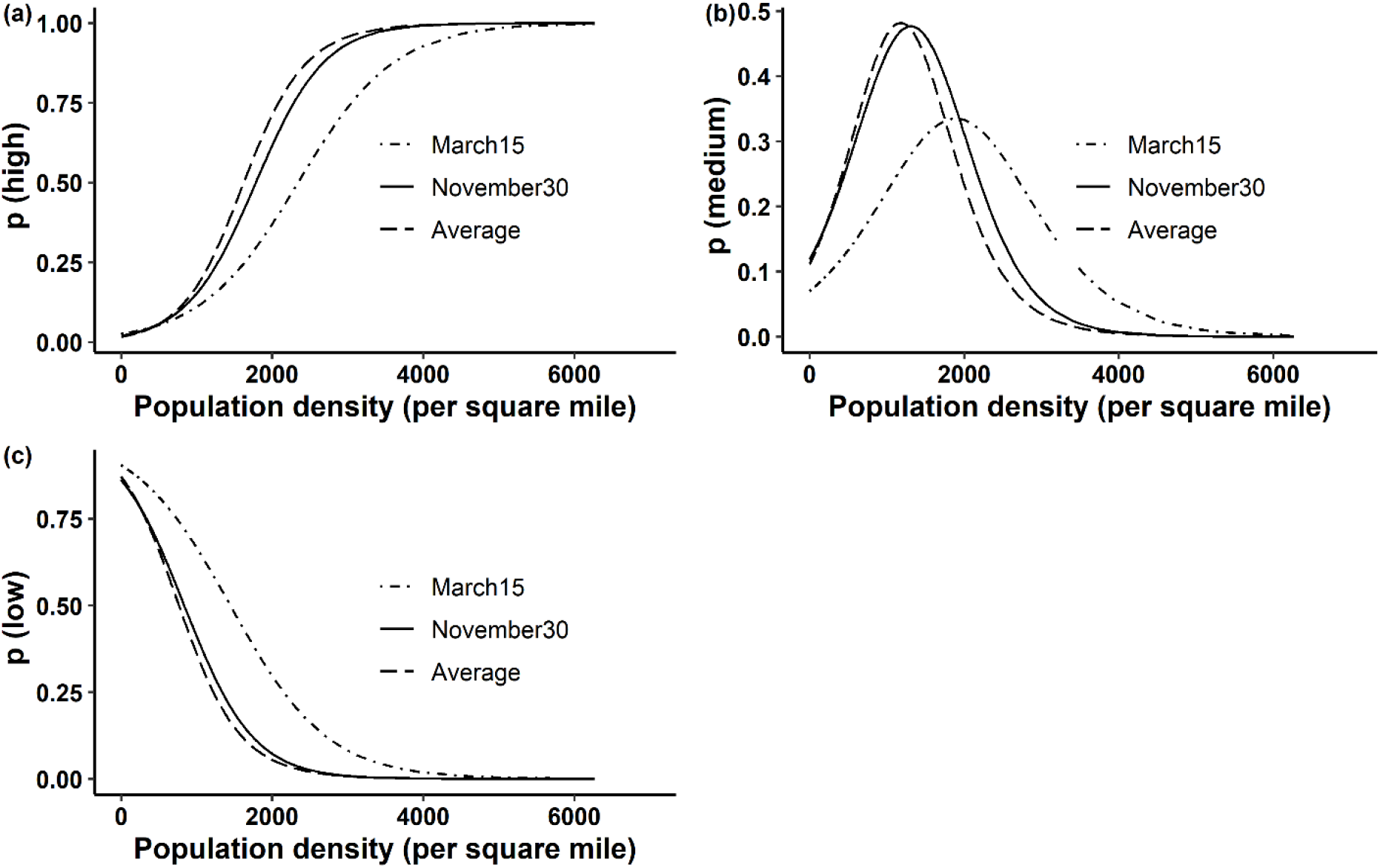
Changes in bi-weekly probability from March 15, 2020, to November 30, 2020 of (a) high, (b) medium, and (c) low number of cases

In Figure 2(a), the threshold population density is shown at which a 50% chance of a high number of cases will occur, ca. 1,622 per square mile. The population density for getting a low number of cases at 50% chance is 762 per square mile (Figure 2(c)), and the arithmetic mean of these two values at high and low cases gives an average of 1,192 per square mile and defined as the population density at which there is a 50% chance of infection. Thus, 4.02% (125 of 3,107) of the counties with population density greater than 1,192 per square mile are at 50% or greater risk of infection as on November 30,2020. The key results discussed here are concisely summarized in Table 3.

**Table 3:**
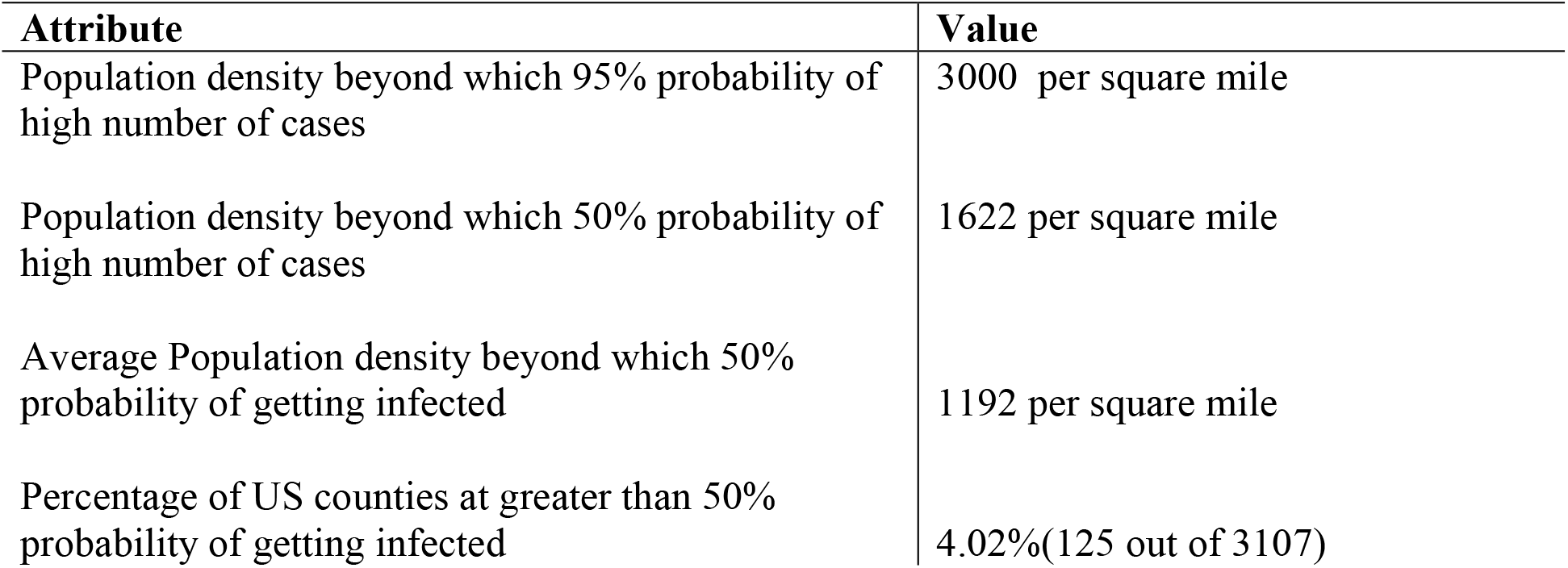
Key results of Population density-cases analysis

Table 4 provides values for population density and arithmetic average probability of high number of cases for each state in the US, as of November 30, 2020.

**Table 4:** Probabilities of high number of COVID19 cases as of November 30, 2020

| State | Population density (per square mile) | Average probability of high cases (Ap) | Maximum probability of high cases (Map) | Minimum probability of high cases (Minp) | Rank of Ap | Rank of Map | Percent difference between Map and Ap |
| --- | --- | --- | --- | --- | --- | --- | --- |
| District of Columbia | 11569.7 | 1 | 1 | 1.000 | 1 | 1 | 0 |
| New Jersey | 1207.8 | 0.404 | 1 | 0.027 | 2 | 1 | 60 |
| Rhode Island | 1024.5 | 0.28 | 0.652 | 0.043 | 3 | 23 | 57 |
| Massachusetts | 883.5 | 0.242 | 1 | 0.022 | 4 | 1 | 76 |
| Connecticut | 736.6 | 0.156 | 0.421 | 0.028 | 7 | 27 | 63 |
| Maryland | 622.9 | 0.165 | 1 | 0.019 | 6 | 1 | 84 |
| Delaware | 499.6 | 0.144 | 0.368 | 0.030 | 8 | 29 | 61 |
| New York | 412.8 | 0.099 | 1 | 0.017 | 11 | 1 | 90 |
| Florida | 400.7 | 0.078 | 0.981 | 0.018 | 13 | 14 | 92 |
| Pennsylvania | 286.1 | 0.079 | 1 | 0.018 | 12 | 1 | 92 |
| Ohio | 286.1 | 0.066 | 0.927 | 0.019 | 14 | 18 | 93 |
| California | 253.7 | 0.12 | 1 | 0.017 | 10 | 1 | 88 |
| Illinois | 228.2 | 0.047 | 0.999 | 0.018 | 18 | 10 | 95 |
| Hawaii | 220.3 | 0.121 | 0.52 | 0.018 | 9 | 26 | 77 |
| Virginia | 216.1 | 0.232 | 1 | 0.018 | 5 | 1 | 77 |
| North Carolina | 215.7 | 0.041 | 0.711 | 0.018 | 20 | 20 | 94 |
| Indiana | 187.9 | 0.038 | 0.857 | 0.018 | 24 | 19 | 96 |
| Georgia | 184.6 | 0.051 | 0.93 | 0.018 | 15 | 17 | 95 |
| Michigan | 176.7 | 0.048 | 0.944 | 0.018 | 17 | 16 | 95 |
| South Carolina | 171.2 | 0.027 | 0.077 | 0.018 | 32 | 35 | 65 |
| Tennessee | 165.6 | 0.031 | 0.346 | 0.018 | 28 | 30 | 91 |
| New Hampshire | 151.9 | 0.029 | 0.053 | 0.018 | 31 | 39 | 45 |
| Washington | 114.6 | 0.03 | 0.181 | 0.017 | 30 | 33 | 83 |
| Kentucky | 113.1 | 0.031 | 0.706 | 0.018 | 28 | 21 | 96 |
| Texas | 111 | 0.036 | 0.952 | 0.017 | 27 | 15 | 96 |
| Louisiana | 107.6 | 0.04 | 0.689 | 0.018 | 22 | 22 | 94 |
| Wisconsin | 107.5 | 0.038 | 0.993 | 0.018 | 24 | 12 | 96 |
| Alabama | 96.8 | 0.022 | 0.071 | 0.018 | 37 | 36 | 69 |
| Missouri | 89.3 | 0.037 | 0.999 | 0.018 | 26 | 10 | 96 |
| West Virginia | 74.6 | 0.023 | 0.046 | 0.018 | 35 | 42 | 50 |
| Minnesota | 70.8 | 0.043 | 0.984 | 0.017 | 19 | 13 | 96 |
| Vermont | 67.7 | 0.021 | 0.036 | 0.018 | 39 | 46 | 42 |
| Arizona | 64.1 | 0.021 | 0.053 | 0.018 | 39 | 39 | 60 |
| Mississippi | 63.4 | 0.02 | 0.042 | 0.017 | 42 | 44 | 52 |
| Arkansas | 58 | 0.02 | 0.059 | 0.018 | 42 | 38 | 66 |
| Oklahoma | 57.7 | 0.025 | 0.225 | 0.017 | 33 | 32 | 89 |
| Iowa | 56.5 | 0.021 | 0.118 | 0.018 | 39 | 34 | 82 |
| Colorado | 55.6 | 0.049 | 1 | 0.017 | 16 | 1 | 95 |
| Oregon | 43.9 | 0.039 | 0.637 | 0.017 | 23 | 24 | 94 |
| Maine | 43.6 | 0.022 | 0.038 | 0.018 | 37 | 45 | 42 |
| Utah | 39 | 0.041 | 0.414 | 0.017 | 20 | 28 | 90 |
| Kansas | 35.6 | 0.023 | 0.272 | 0.017 | 35 | 31 | 92 |
| Nevada | 28.1 | 0.02 | 0.043 | 0.017 | 42 | 43 | 53 |
| Nebraska | 25.2 | 0.025 | 0.527 | 0.017 | 33 | 25 | 95 |
| Idaho | 21.6 | 0.019 | 0.048 | 0.017 | 46 | 41 | 60 |
| New Mexico | 17.3 | 0.02 | 0.069 | 0.017 | 42 | 37 | 71 |
| South Dakota | 11.7 | 0.018 | 0.03 | 0.017 | 47 | 47 | 40 |
| North Dakota | 11 | 0.018 | 0.022 | 0.017 | 47 | 48 | 18 |
| Montana | 7.3 | 0.018 | 0.02 | 0.017 | 47 | 49 | 10 |
| Wyoming | 6 | 0.018 | 0.019 | 0.017 | 47 | 50 | 5 |

This average probability is the simple arithmetic average of the respective probabilities of state counties. It is intuitive that the most densely populated states were also those with the highest probability of a high number of cases, strengthening the finding that population density is critical beyond a specific threshold. The average probability for a high number of COVID-19 cases provides a number useful for conceptualizing the overall risk of infection in a particular state. However, except for a few densely populated states, epicenter counties are not highlighted. For example, on November 30, 2020, Texas, California, Florida, and Illinois were States with the largest number of COVID-19 cases. From Table 4, it was obvious that the relatively low values of average probabilities for those four states do not reflect their epicenter status. Therefore, we defined the maximum average probability for a state which is taken equal to the maximum value of average probabilities considering all the counties of a state. Thus, the maximum average probability for each state as calculated. The high probability of a high number of cases (>90%) indicated this metric performed acceptably to rank a state as an epicenter. Exceptions were noted, where relatively low value of maximum average probability of high cases was observed in low population density states having high number of COVID-19 cases. This anomaly is a potential limitation of the logistical regression methods, in dealing with locations with low population densities and high cases. However, it is understood that any regressive model considering population density as the single explanatory variable likely would fail to explain a high number of cases in less densely populated regions. Lastly, an important observation with reference to population density and case analysis can be discerned from Table 4, namely the percentage difference between the maximum and average probabilities of high number of cases, being very high for many states, notably those with high maximum average probabilities. This signifies that only a few counties of the state account for a large number of cases, and the state as a whole would not be an epicenter of COVID-19.

The analysis in our study assumes a uniform population distribution. In reality, the population is generally not distributed evenly across the county, as most of the population clusters in and around cities. The lack of a standard sub-county level case count, which rules out the possibility of conducting a more realistic city-level threshold analysis, forms a limitation of our study.Furthermore, though county population density is generally considered a reliable predictor to explain COVID-19 cases due to its high explanatory power (Riley, 2007; Wong & Li, 2020), controlling it for other variables such as population size could bring valuable insights. to determine running thresholds on population density.

## 4. Conclusions and Implications on Future of urban cities

The interrelationship of population density with the number of COVID-19 cases was analyzed, with the objective to determine thresholds for population density above which there was a 50% or greater risk of COVID-19 cases in humans. Population density and COVID-19 cases, when analyzed together, suggest *ca*. 4% of the counties (shown in Figure 4) in the United States would be at 50% or greater at risk of COVID-19 cases and confined to a few counties.

**Figure 4:**
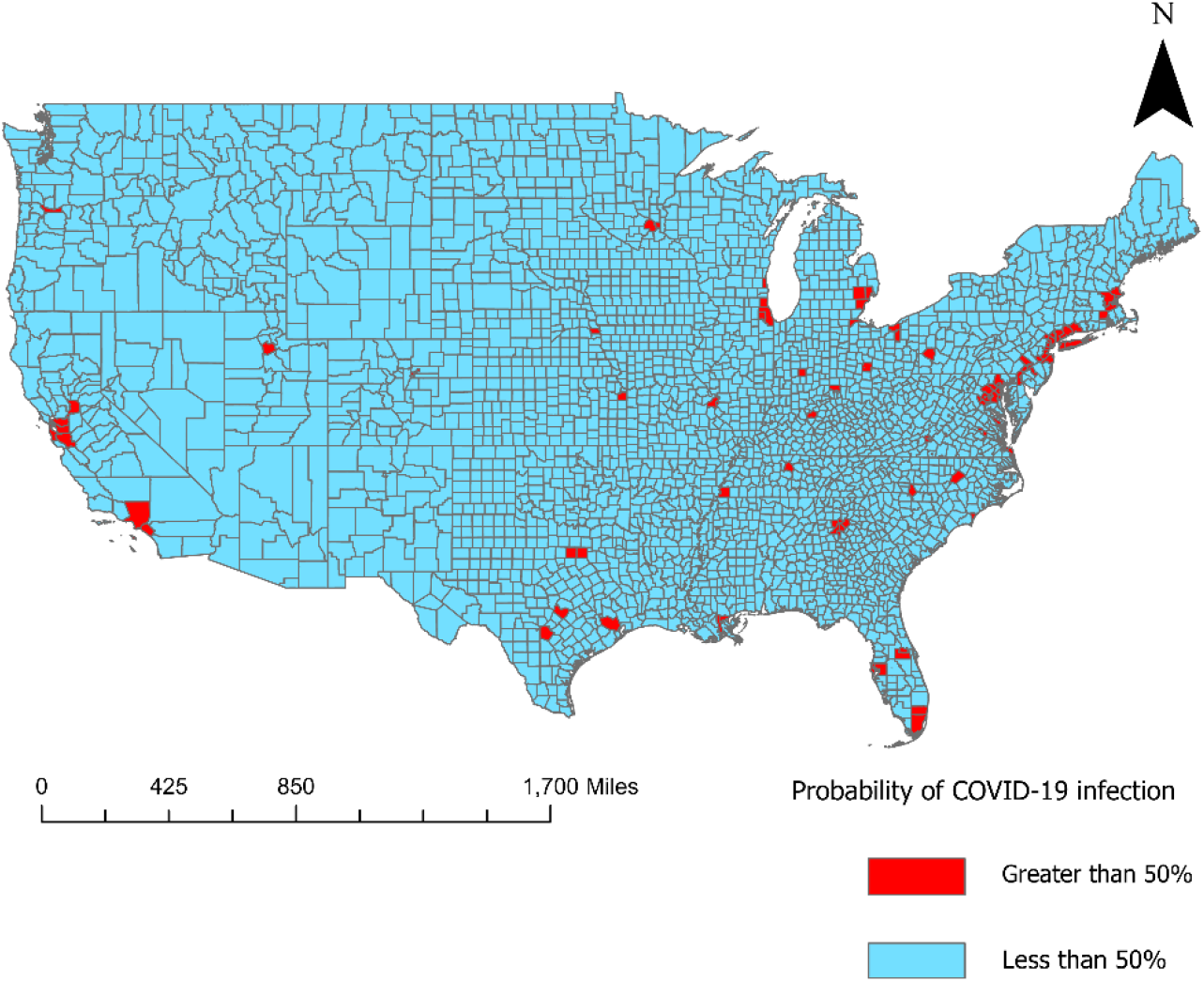
US counties with greater than 50% probability of getting infected as of November 30,2020.

The thresholds provide useful information as a guide for policymakers. In combination with other governing factors, the population density threshold can provide a more decisive conclusion, notably for estimating cases and mitigating COVID-19 in human cases, especially for urban neighborhoods that are more likely heterogenous in race, income, and infrastructure (J. A. Maantay, Maroko, & Herrmann, 2007; J. Maantay & Maroko, 2009). Dense populations comprise sub-populations, namely communities of color and low-income communities that are vulnerable, e.g. poor housing, high pollution, lack of access to health care, and a higher rate of pre-existing conditions(Brulle & Pellow, 2006; Bullard, 2005; Pellow, 2000). Nevertheless, the relationship between population density and rates of infection is sufficiently robust that it can be employed by policymakers to prepare anticipatory plans for specific communities and thereby prevent the spread of infection and mitigate the effects of the disease.

## Data Availability

Raw data sets are publicly available and can be accessed using weblinks provided.
Datasets generated in this study are available on openly accessible data servers.

https://github.com/nytimes/covid-19-data

https://www.census.gov/data/datasets/time-series/demo/popest/2010s-counties-total.html

https://www.census.gov/library/publications/2011/compendia/usa-counties-2011.html#LND

## Data Availability

Raw data sets are publicly available and can be accessed using weblinks provided. Datasets generated in this study are available on openly accessible data servers.

https://github.com/nytimes/covid-19-data

https://www.census.gov/data/datasets/time-series/demo/popest/2010s-counties-total.html

https://www.census.gov/library/publications/2011/compendia/usa-counties-2011.html#LND

While there are many other links to get the data on COVID-19 case numbers, to the best of our knowledge, GitHub is the only concise yet comprehensive source which provides easy to analyze chronological case count data for US at county scale.

## Code Availability

All the data analysis was performed using the MINITAB software package, a standard package for statistical analysis available at https://www.minitab.com/en-us/products/minitab/

## Author Contributions

A.J. is responsible for the main concepts; Y.J. and M.G. wrote the study; Y.J., M.G., M.U. and A.J. carried out the analyses; C.Y.W., T.N. A.A. and R.C. prepared the final manuscript.

## Corresponding author

Antarpreet Jutla

## List of tables

Table S1: Results from the ordinal logistic regression model (November 15,2020)

