## Supplementary file for "Identification of thresholds on population density for understanding transmission of COVID-19"

### Supplementary section: methods

#### Theory of ordinal logistic regression

For any explanatory variable (covariate) *X* for response variable Y, a simple linear regression model linearly relates *Y* to *X*.

$Y = \alpha + \beta X$ [1]

The simple linear regression fails when this assumption does not hold, and nonlinear regression should be considered. For example, the simple linear regression model is invalid when *Y* is binary because linear regression assumes that *Y* can take any numeric value between minus infinity and infinity. The relationship between *Y* and *X* can be described by modeling the probability ,p,of an event instead of *Y* itself,

$p = \mathrm{P}(Y = 1)$ [2]

*Y* can be 0 or 1, and p can be any value between 0 and 1.

Odds (or odds ratio) will be $\frac{p}{1-p}$ and can be the positive value, and the logit (logarithmic of odds) can be any positive or negative value. Therefore, a linear relationship can be assumed between the logit and *X*:

$$logit = \ln\left(\frac{p}{1-p}\right) = \alpha + \beta X \quad [3]$$

Also,

$$p = \frac{\exp\ (\alpha+\beta X)}{1+\exp\ (\alpha+\beta X)} \quad [4]$$

This model is called the logistic regression model (Hosmer& Lemeshow , 2000,p.6). Occasionally the response variable *Y* can have more than two values (0 and 1) and follow order i.e., ordinal. Simple logistic regression cannot be applied; hence cumulative probabilities, cumulative odds and cumulative logits are used.

For k+1 ordered categories, $P(Y \ <= \ \mathrm{i}) = \ \mathrm{p1} \ + \ \mathrm{p2} \ + \ \ldots..\mathrm{pi}$

$$logit = \ln\left(\frac{P(Y\leq i)}{1-P(Y\leq i)}\right) \qquad , i = 1,2,\ldots,k \quad [5]$$

Hence, for *m* covariates the cumulative logistic model for ordinal response data is

$$logit\ (Y \leq i) = \alpha + \beta_{i1}X + \cdots + \beta_{im}X_m \quad [6]$$

The general cumulative logistic regression model contains many parameters as we have k model equations and one logistic coefficient $\beta_{ij}$ for each covariate combination.

When the ordinal response *Y* is related to a continuous variable and it forms the proportional odds model where the logistic coefficients do not depend on *i*, we have only one common parameter βj for each covariate. The cumulative odds are

$$odds\ (Y \leq i) = \exp(\alpha_i) + \exp(\beta_1 X_1 + \cdots + \beta_m X_m) \qquad , i = 1,2,\ldots,k \ [7]$$

It signifies that the odds differ only for the intercepts α, for each cut-off category, i.e. the odds are proportional. MINITAB uses the proportional odds model where a vector of predictors, *X*, has a parameter β describing the effect of *X* on the log odds of the response in category *k* or below. Minitab assumes an identical effect of x for all *k* – 1 category, so only one coefficient is calculated for each predictor. The coefficient for the predictor indicates that for any fixed *k*, the estimated change in the logit of the response when predictor is at one level compared to the reference level. Here in this study, the proportional odds model is used where response *Y* (COVID19 cases) is a categorical variable, grouped as low, medium, and high cases, and a vector of the predictor (population density), *X*, has a parameter β describing the effect of *X* on the log odds of the response *Y* in category *k* or below.

To determine whether the pairs are concordant or discordant, MINITAB calculates the cumulative predicted probabilities of each observation and compares these values for each pair of observations.

**Concordant pairs**

For pairs that include the lowest response value (in the example above, that is 1), a pair is concordant if the cumulative probability up to the lowest response value is greater for the observation with the lowest response value than for the observation with the higher response value. For pairs with the highest response values (in the example above, pairs with 2 and 3), a pair is concordant if the cumulative probability up to 2 is greater for the observation with the response value 2 than the observation with the response value 3.

**Discordant pairs**

For pairs that include the lowest response value (in the example above, that is 1), a pair is discordant if the cumulative probability up to the lowest response value is greater for the observation with the higher response value than for the observation with the lower response value. For pairs with the highest response values (in the example above, pairs with 2 and 3), a pair is discordant if the cumulative probability up to 2 is greater for the observation with the response value 3 than the observation with the response value 2.

**Ties**

A pair is tied if the observations have equal cumulative probabilities.

The below mentioned terms explain the key components of the fitted ordinal logistic model for testing its adequacy and subsequently for interpretation of the results

i. **Link function**

In general, a link function (Hosmer& Lemeshow ,2000,p.48) is required in logistic regression to express the dependent variable as a linear function of the independent variables. In this study, it relates the response(ordinal cases) to linear predictor (population density). It transforms the probabilities of ordinal response to the continuous scale of [0,1].The present study uses a widely used logit link function (Hosmer& Lemeshow , 2000,p.48) .The general form of the link function can be written as:

$g(x_k) = \theta_k + y'\beta, k = 1, \dots \dots \dots K - 1$ [8]

Where $g(x_k)$ is the logit link function given by $g(x) = log_e(\frac{p}{1-p})$

p is the event probability

K=number of distinct categories of the response (three (03) in our study, where high=1;medium=2,and low=3)

$\theta_k$ =constant associated with the k$^{th}$ distinct response category

$y$ = a vector of predictor variables

$\beta$= a vector of coefficients associated with the predictors

#### ii. Coefficients and constants

Coefficients are used to examine the change in the probability of response variable due to the change in predictor variables. A positive value indicates that an increase in predictor value more likely than otherwise. Negative values signify that the last event or the event close to it is more likely than otherwise. A value close to zero signifies that predictor has a little effect on the response probability. Proportional odds model was used, and it assumes that the effects of predictor variables are the same for all categories on the logarithmic scale. It further implies that the model has different intercepts but common slopes (coefficients) among categories. Thus, the general form of regression equation using logit function is given by

$$ln\left(\frac{p(event_j)}{1-p(event_j)}\right) = \alpha_j + \beta_1 X_1 + \beta_2 X_2 + \cdots \beta_n X_n \quad [9]$$

where, *p(event)* is the cumulative probability of occurrence of categorical response category *j*. The constants and coefficients are denoted by $\alpha_j$ and $\beta = [\beta_1\ \beta_2 \ldots \beta_n]$ respectively.

For this study, the regression equations were

$$ln\left(\frac{p(high\ cases)}{1-p(high\ cases)}\right) = \alpha_1 + \beta * population\ density \ [10]$$

*1*

$$ln\left(\frac{p(medium\ cases)}{1-p(medium\ cases)}\right) = \alpha_2 + \beta * population\ density \ [11]$$

If the number of categories were *j*, then *j*-1 regression equations suffice to determine the cumulative probabilities for each of the j categories as the probabilities for $j^{th}$ category was 1.

In Table S1, constant 1 (high cases) and constant 2 (medium cases) denote the values of $\alpha_1$ and $\alpha_2$ respectively, while the coefficient for population density is denoted by $\beta$. These constants and coefficients were estimated using maximum likelihood estimation method (Hosmer & Lemeshow, 2000,p.63).

**iii. Standard error of coefficients**

It represents the precision. A smaller value indicates greater precision. ("Methods and formulas for Ordinal Logistic Regression - Minitab," 2019)

**iv. Z-value**

Z-value statistic is the ratio of coefficient(or constant) to its standard error. A large value indicates significant relationship between the predictor and response. ("Methods and formulas for Ordinal Logistic Regression - Minitab," 2019)

**v. p- value**

p-values measures statistical significance. A value less than the chosen significance level is generally sufficient to reject the null hypothesis. A significance level of 0.05 is generally used which means that there is 5% risk of concluding that relationship exists between the response and model terms. In this study, p-values for each of the constants and coefficient were zero at significance level of 0.05 (Table S1) signifying that a significant association exists between the response and the terms (constants and coefficients).

**vi. Odds ratio**

Odds ratio is used to understand the effect of predictor on the response probability. Mathematically defined as

$$odds\ ratio = \frac{probability\ of\ event\ happening}{probability\ of\ event\ not\ happening}$$

A value greater than 1 indicates that the with the increase in predictor, first event and the events closer to the first event become more likely.

Within the context of this study, value of odds ratio is greater than 1 implies that the probability of high cases increases with the increase in population density.

**vii. Log-likelihood**

This metric is mainly used to compare two models to help in model selection. A value close to zero suggest that the data fits the model well (Hosmer & Lemeshow, 2000,p.9).

**viii. Test of all slopes equal to zero**

This test is used to determine whether at least one of the predictors in the model has a statistically significant association with the response events. Degree of Freedom (DF) is the number of coefficients of predictors used in the model. A p-value less than or equal to the significance level(usually 0.05) indicates that statistically significant association exists between the response and at least one of the predictors("Methods and formulas for Ordinal Logistic Regression - Minitab," 2019)

.

**ix. Deviance goodness of fit test**

A statistic that indicates that how well the model fits the data. A p-value greater than 0.05 indicates good fit (Hosmer & Lemeshow, 2000,p.37).

**x. Measures of association**

Concordant and discordant pairs

Concordant and discordant pairs indicate how well the model predicts data. The more concordant pairs imply greater predictive ability of the model ("Methods and formulas for Ordinal Logistic Regression - Minitab," 2019).Key statistical measures based on Concordant and discordant pairs are as below:

**Somer's D**

Somers' D is the percentage difference between concordant and discordant pairs, including ties.

**Goodman-Kruskal Gamma**

Goodman-Kruskal Gamma is the percentage difference between concordant and discordant pairs, excluding ties.

**Kendall's Tau-a**

Kendall's Tau-a is the percentage difference of concordant and discordant pairs out of all possible pairs, including pairs with the same response value.

All the three above mentioned measures of association are used to compare the predictive performance of the models. They can be estimated by formulas given below ("Methods and formulas for Ordinal Logistic Regression - Minitab," 2019).

$$Somer's\ D = \frac{C - D}{C + D + T}$$

$$Goodman - Kruskal\ Gamma = \frac{C - D}{C + D}$$

$$Kendall's\ Tau - a = \frac{C - D}{0.5 * N * (N - 1)}$$

Where, C=number of concordant pairs

D=number of discordant pairs

T= number of tied pairs

N=Total number of observations

**Table S1**: Sample output from the model for November 15, 2020.

<table>
<tr><td colspan="6">1.link function: logit</td></tr>
<tr><td colspan="6"></td></tr>
<tr><td colspan="6">2.Response information</td></tr>
<tr><td rowspan="5" colspan="2">Levels_Nov30</td><td colspan="2">value</td><td colspan="2">count</td></tr>
<tr><td colspan="2">high</td><td colspan="2">155</td></tr>
<tr><td colspan="2">medium</td><td colspan="2">464</td></tr>
<tr><td colspan="2">low</td><td colspan="2">2488</td></tr>
<tr><td colspan="2">Total</td><td colspan="2">3107</td></tr>
<tr><td colspan="6"></td></tr>
<tr><td colspan="6">3.Logistic regression table</td></tr>
<tr><td>predictor</td><td>coefficient</td><td colspan="2">Standard error of coefficient</td><td>Z-Value</td><td>p-value</td></tr>
<tr><td>Constant1(high)</td><td>-3.90253</td><td colspan="2">0.115654</td><td>-33.74</td><td>0</td></tr>
<tr><td>Constant2 (medium)</td><td>-1.82834</td><td colspan="2">0.0538937</td><td>-33.94</td><td>0</td></tr>
<tr><td>Population density</td><td>0.0021885</td><td colspan="2">0.0001187</td><td>18.44</td><td>0</td></tr>
<tr><td colspan="6"></td></tr>
<tr><td>Predictor</td><td>Odds ratio</td><td colspan="4">95% confidence intervals</td></tr>
<tr><td rowspan="2">Population density</td><td rowspan="2">1.00219</td><td colspan="2">Lower bound</td><td colspan="2">Upper bound</td></tr>
<tr><td colspan="2">1.00196</td><td colspan="2">1.00242</td></tr>
<tr><td colspan="6"></td></tr>
</table>

4.log-likelihood=-1630.720

5.Test of all slopes equal to zero

| DF | G | p-value |
|---|---|---|
| 1 | 538.115 | 0 |

6.Goodness of fit test

| **Method** | **Chi-square** | **DF** | **p-value** |
|---|---|---|---|
| **Deviance** | 1.0021E+12 | 6211 | 1.000 |

7.Measures of association (between the response variable and predicted probabilities)

| **Pairs** | **Number** | **Percent** | **Summary measures** | **value** |
|---|---|---|---|---|
| Concordant | 1477335 | 91.6 | Somer’s D | 0.84 |
| Discordant | 117843 | 7.3 | Goodman-Kruskal Gamma | 0.85 |
| Ties | 16814 | 1.0 | Kendall’s Tau | 0.28 |
| **Total** | **1611992** | **100.0** | | |
